# Hesitant or not? A global survey of potential acceptance of a COVID-19 vaccine

**DOI:** 10.1101/2020.08.23.20180307

**Authors:** Jeffrey V Lazarus, Scott Ratzan, Adam Palayew, Lawrence O Gostin, Heidi J Larson, Kenneth Rabin, Spencer Kimball, Ayman El-Mohandes

## Abstract

A number of COVID-19 vaccines are under development, with one or more possibly becoming available in 2021. We conducted a global survey in June 2020 of 13,426 people in 19 countries to determine potential acceptance rates of a COVID-19 vaccine and factors influencing acceptance. We ran univariate logistic regressions to examine the associations with demographic variables. 71.5% reported they would be very or somewhat likely to take a COVID-19 vaccine; 61.4% reported they would accept their employer’s recommendation to take a COVID-19 vaccine. Differences in acceptance across countries ranged from almost 9 in 10 (China) to fewer than 6 in 10 (Russia). Respondents reporting higher levels of trust in information from government sources were more likely to accept a vaccine, and take their employer’s advice to do so. Targeted interventions addressing age, sex, income, and education level are required to increase and sustain public acceptance of a COVID-19 vaccine.

---

The COVID-19 pandemic is expected to continue to impose enormous burdens of morbidity and mortality while severely disrupting societies and economies worldwide. Governments must be ready to ensure large-scale equitable access and distribution once safe and effective vaccines become available. This requires sufficient health system capacity and strategies to enhance acceptance of, and trust in, the vaccine and its delivery. In many countries, vaccine hesitancy and misinformation present substantial obstacles to achieving coverage and community immunity^1,2^.

In 2015, the World Health Organization (WHO)-hosted Strategic Advisory Group of Experts (SAGE) on Immunization defined vaccine hesitancy as “delay in acceptance or refusal of vaccination despite availability of vaccination services”, and that it…” is complex and context-specific, varying across time, place, and vaccines”^3^, as has been confirmed in multiple studies^4,5^. Concern about vaccine hesitancy is growing worldwide^6^; WHO identified vaccine hesitancy as one of the top ten global health threats in 2019^7^.

Governments, public health officials, and advocacy groups must be prepared to address hesitancy, should a COVID-19 vaccine become available. Anti-vaccination activists are already campaigning in multiple countries against the need for a vaccine, with some denying the existence of COVID-19. Misinformation spread through multiple channels could have a considerable impact on the acceptance of a COVID-19 vaccine^8^. The accelerated pace of vaccine development has further heightened public anxieties, and could compromise acceptance^9^.

Governments and societies must gauge current levels of willingness to receive a potentially safe and effective COVID-19 vaccine, and identify correlates of vaccine hesitancy/acceptance. We present findings from a study on the likelihood of vaccine acceptance from a sample of 13 426 respondents in 19 countries.

## Methods

We analysed two questions from the COVID-SCORE study pertaining to COVID-19 vaccine acceptance^10^. In that survey, participants responded to a total of 22 items, including two related to vaccine uptake, one related to trust in pandemic information sources, and standard demographic questions regarding age, gender, level of education, and household income (*see* Supplementary materials 1).

### Study participants

Participants were recruited by Emerson College Polling through international online panel providers: Dynata provided 7423 respondents across all 19 countries; Opinion Access provided 3293 respondents from 14 countries; Survey Monkey provided 1941 responses from 12 countries; and Amazon MTurk provided 762 respondents from eight countries. Respondents’ identities were verified using IP addresses and mobile phone numbers to ensure that each participant was real and unique upon initial registration. Participants were recruited for the panels via a variety of methods, including online, telephone, and direct mail solicitation. Sampling was random and is described in detail elsewhere^10^.

### Data collection

Survey data were collected from 16 to 20 June 2020 from an online panel of 13 426 respondents aged 18 years and older from 19 countries from among the top 35 impacted by the pandemic, ranging between 619 and 773 participants per country. To ensure regional representation, we selected the next most affected country from regions not represented on the top 35 list: Brazil, Canada, China, Ecuador, France, Germany, India, Italy, Mexico, Nigeria, Poland, Russia, Singapore, South Africa, South Korea, Spain, Sweden, the United Kingdom, and the United States.

The more general vaccine-related question was, “If a COVID-19 vaccine is proven safe and effective and is available to me, I will take it.” Respondents were also asked to register their level of agreement with a second statement: “I would follow my employer’s recommendation to get a COVID-19 vaccine once the government has approved it as safe and effective.” Responses were recorded on a 5-point Likert scale (“completely disagree,” “somewhat disagree,” “neutral/no opinion,” “somewhat agree,” “completely agree”). We examined the demographic breakdown of the responses to these questions. Data for age and income were collected through open-text fields. Age was coded into age categories: 18–24, 25–54, 55–64 and 65 years and older. Where respondents provided income information, the levels were categorized as “<$2 a day”, “$2–$8 a day”, “$8–$32 a day”, and “+$32”. Education levels were categorized as less than high school (low), high school or some college (medium), bachelor’s degree (high), and postgraduate (very high). Gender was defined as male, female or other. We also collected information on whether the respondent or a family member had been sick with COVID-19, and COVID-19 cases and deaths per million population at the country level^11^. For cases per million population and mortality per million population, we categorized the continuous values into categories of low, medium, and high. For cases per million population, low was < 2000 cases per million population, medium was between 2000 and 4000 cases per million population, and high was greater than 4000 cases per million population. For mortality per million population, low was defined as < 200 deaths per million population, medium as between 200 and 400 deaths per million population, and high as > 400 deaths per million population.

### Analysis

We analysed the distribution of the responses against the different questions for the entire dataset and further examined differences by country. We calculated results for two sets of univariate regressions: one for each of the two questions related to vaccines. We used logistic regression, defining the outcome as 1 if a respondent answered, “completely agree” or “somewhat agree” and 0 for any other response. The independent demographic variables were: age, gender, income, and education. We also examined the relationship between the two regression outcomes and whether someone in the respondent’s family had been sick with COVID-19, as well as existing country-by-country data on COVID-19 cases per million population, COVID-19 mortality per million population, and whether a respondent reported that they trusted pandemic information from their government (yes/no).

## Results

The 13 426 respondents from 19 countries represented 55% of the world population (Table 1). Women comprised 53.5%, and 63.3% earned above $32 dollars. Most respondents (36.4%) had a university degree, and 62.4% were between 25 and 54 years (Table 1).

**Table 1.** Description of participants and breakdown of the two COVID-19 vaccine questions.

|  | Overall |
| --- | --- |
| N | 13 426 |
| Gender (%) |  |
| Female | 7 172 (53.5) |
| Male | 6 129 (45.8) |
| Other | 94 (0.7) |
| Gapminder income level (%) |  |
| <US\$ 2 per day | 447 (3.3) |
| \$2–\$8 per day | 840 (6.3) |
| \$8–\$32 per day | 3 011 (22.4) |
| \$32+ | 8 498 (63.3) |
| Did not answer | 630 (4.7) |
| Education level (%) |  |
| Less than high school | 3 830 (28.6) |
| High school, some college | 4 692 (35.0) |
| Bachelor | 3 694 (27.6) |
| Postgraduate | 1 179 (8.8) |
| Age group in years (%) |  |
| 18–24 | 2 057 (15.4) |
| 25–54 | 8 360 (62.4) |
| 55–64 | 1 493 (11.1) |
| 65+ | 1 485 (11.1) |
| Accept COVID-19 vaccine if generally available (%) |  |
| Completely agree | 6 288 (46.8) |
| Somewhat agree | 3 316 (24.7) |
| Neutral/no opinion | 1 912 (14.2) |
| Somewhat disagree | 819 (6.1) |
| Completely disagree | 1 091 (8.1) |
| Accept COVID-19 vaccine if employer recommended it (%) |  |
| Completely agree | 4 286 (31.9) |
| Somewhat agree | 3 957 (29.5) |
| Neutral/no opinion | 2 772 (20.6) |
| Somewhat disagree | 1 090 (8.1) |
| Completely disagree | 1 321 (9.8) |

Characteristics of respondents’ and their answers to whether they would take a “proven safe and effective” COVID-19 vaccine are listed in Table 1. China reported the highest proportion of positive responses (88.6%) and the lowest proportion of negative responses (0.7%); while Poland reported the highest proportion of negative responses (27.3%) and Russia the lowest proportion of positive responses (58.9%). China also had the highest proportion of positive responses (86.2%) and the lowest proportion of negative responses (0.7%) to the question on whether they would accept the vaccine if recommended by their employer, while Russia had the highest proportion of negative responses and the lowest proportion of favorable responses (Supp Table 1).

When asked if, “You would accept a vaccine if it were recommended by your employer and was approved safe and effective by the government”, 31.9% completely agreed, while 17.9% somewhat or completely disagreed (Table 1). There was considerable variation by country, with China having the highest proportion of positive responses (86.2%) and the lowest proportion of negative responses(0.7%). Russia had the highest proportion of negative responses (27.9%) and the lowest proportion of respondents (46.7%) willing to accept their employer’s recommendation (Supp Table 1).

We report results for the 16 regressions in Table 2. Older people were more likely to accept the vaccine. This difference was strongest (OR 1.73) when comparing the oldest to the youngest age cohort (Table 2). The opposite trend was observed for accepting the vaccine if one’s employer required it. Gender differences were small, but the univariate association for both questions suggested that men were slightly less likely to respond positively than women.

**Table 2:** Univariate regression outputs for vaccine acceptability questions against demographics and variables of interest.

|  | Beta-coefficients of vaccine questions<br>(95% confidence intervals) |  | Beta-coefficients of business question<br>(95% confidence intervals) |  |
| --- | --- | --- | --- | --- |
| Age (years) | 25–55 vs 18–24 | 1.12 (1.01, 1.25) | 25–55 vs 18–24 | 0.95 (0.86, 1.05) |
|  | 55–64 vs 18–24 | 1.21 (1.04, 1.40) | 55–64 vs 18–24 | 0.84 (0.73, 0.96) |
|  | 65 + vs 18–24 | 1.73 (1.48, 2.02) | 65 + vs 18–24 | 0.78 (0.68, 0.89) |
| Sex | Male vs female | 0.84 (0.78, 0.91) | Male vs female | 0.87 (0.81, 0.93) |
|  | Other vs female | 0.22 (0.14, 0.33) | Other vs female | 0.32 (0.21, 0.49) |
| Income | \$2–\$8 vs <\$2 | 1.38 (1.09, 1.74) | \$2–8 vs <\$ | 0.91 (0.72, 1.14) |
| | \$8–\$32 vs <\$2 | 1.87 (1.53, 2.29) | \$8–32 vs <\$2 | 1.04 (0.85, 1.27) |
| | \$32+ vs <\$2 | 2.18 (1.79, 2.64) | \$32+ vs <\$2 | 1.47 (1.21, 1.79) |
| | refused vs <\$2 | 0.91 (0.71, 1.16) | refused vs <\$2 | 0.78 (0.61, 1.00) |
| Education | Medium vs low | 1.26 (1.15, 1.39) | Medium vs low | 1.26 (1.15, 1.37) |
|  | High vs low | 1.34 (1.21, 1.48) | High vs low | 1.24 (1.13, 1.36) |
|  | Very high vs low | 1.45 (1.25, 1.69) | Very high vs low | 1.31 (1.15, 1.49) |
| Myself or family sick with COVID | Yes vs no | 0.97 (0.87, 1.08) | Yes vs no | 1.05 (0.96, 1.71) |
| Cases per million population | Middle vs low | 1.60 (1.46, 1.75) | Middle vs low | 1.30 (1.20, 1.42) |
|  | High vs low | 1.55 (1.42, 1.71) | High vs low | 1.62 (1.49, 1.76) |
| Mortality per million population | Middle vs low | 1.38 (1.25, 1.52) | Middle vs low | 1.25 (1.15, 1.37) |
|  | High vs low | 1.43 (1.30, 1.56) | High vs low | 1.28 (1.18, 1.39) |
| Trust in government | Yes vs no | 1.67 (1.54, 1.80) | Yes vs no | 2.34 (2.20, 2.56) |

People earning above $32/day were 2.18 times more likely to respond positively to the general question compared to those earning less than $2/day. Higher levels of education were associated positively with vaccine acceptance on both questions. People who reported COVID-19 sickness in themselves or family members were no more likely to respond positively to the vaccine question than other respondents. However, cases and mortality per million of a nation’s population were independently associated with a higher likelihood of vaccine acceptance in countries with medium and high disease incidence and mortality.

Respondents who said that they trusted their government were more likely to accept a vaccine compared to those who said that they did not. Moreover, if someone trusted their government, they were more likely to respond positively to their employer’s vaccine recommendation than someone who did not (Table 2).

## Discussion

We conducted a study of potential acceptance of a COVID-19 vaccine in 13 426 randomly selected individuals across 19 high COVID-19 burden countries. Of these, 71.5% responded that they would take a vaccine if it were proven safe and effective, and 61.4% said that they would get vaccinated if their employer recommended it. These numbers varied substantially between countries.

The far-from-universal willingness to accept a COVID-19 vaccine is a cause for concern. Countries where acceptance exceeded 80% tended to be Asian nations with strong trust in central governments (China, South Korea, and Singapore). A relatively high tendency towards acceptance in middle-income countries such as Brazil, India and South Africa is encouraging. Unless and until the origins of such wide variation in willingness to accept a vaccine is better understood and addressed, differences in COVID-19 vaccine coverage between countries could potentially delay the restoration of global connectivity and global economic recovery.

An important finding was the variation across demographically defined groups, being least among those with lower education and income levels. Future vaccine communication interventions should consider the level of scientific and general literacy in sub-populations, identify locally trusted sources of information^12^, and go beyond public announcements that vaccines are safe and effective, and directly address community-specific concerns or misconceptions, historic issues breeding distrust, and be sensitive to predominant religious or philosophical beliefs^13^. Researchers have identified promising interventions for building confidence and reducing vaccine hesitancy in different contexts^14,^15^^ but translating this evidence into large-scale vaccination campaigns will require particular awareness of and attention to existing public perceptions and felt needs. Engaging formal and informal opinion leaders within these communities will be key.

Additionally, we observed age-related associations. Older people were more likely to report that they would take a vaccine, whereas younger respondents were more likely to accept an employer’s vaccine recommendation. Men were less likely than women to accept vaccines in general, or their employer’s recommendation to get vaccinated; however, this association was not large. Those with a higher income were most likely to accept a vaccine. This information may help governments, policymakers, health professionals, and international organizations to more effectively target their messaging around COVID-19 vaccination.

The other source of concern was a discrepancy between reported acceptance of a COVID-19 vaccine and acceptance if vaccination was advocated by one’s employer. All respondents, regardless of nationality, reported they would be less likely to accept a COVID-19 vaccine if it was mandated by employers. This finding across all countries with both high and low reported vaccine acceptance proportions suggests that promoting voluntary acceptance is a better option for employers. It may seem easier to monitor compliance among adults in the working age group if employers required it, but this could fail if it is perceived as limiting employees’ freedom of choice or a manifestation of employers’ self-interest^16^.

A careful balance is required between educating and explaining the necessity of universal vaccine coverage and avoiding a suggestion of coercion. The role of community-based groups that are considered to be impartial may be essential to help build trust in a future COVID-19 vaccine.

Arguably, trust is an intrinsic, and potentially modifiable, component of successful uptake of a COVID-19 vaccine. Our findings show that trust in government, reflecting perceived accountability, is strongly associated with vaccine acceptance and can contribute to public compliance with recommended actions.^17^ Lessons learned from previous infectious disease outbreaks and public health emergencies, including HIV, H1N1, SARS, MERS, and Ebola, remind us that trusted sources of information and guidance are key to disease control^18^.

Clear and consistent communication by government officials is crucial to building public confidence. Credible and culturally informed health communication is vital in influencing positive health behaviors^19,20^, as has been observed with respect to encouraging people to cooperate with COVID-19 control measures.

This survey was conducted in a highly dynamic and changing landscape. At a time when perceived disease threat is higher or lower, it could generate different results. Some of the variables such as the “no response” variable for income and the “other” category for gender had a small number of respondents, and thus that the results may be sensitive to sampling bias.

## Conclusion

In most of the 19 countries surveyed, willingness to accept a COVID-19 vaccine is insufficient. To build greater trust among the population, transparent, evidence-informed policy, and clear communication are needed from stakeholders, and regular feedback from the community. This pandemic provides an important opportunity to build vaccine literacy and confidence to support the uptake of a potential COVID-19 vaccine as well as the overall immunization programme. Efforts must be scaled up now in preparation for the still-elusive promise of a safe and effective COVID-19 vaccine.

## Data Availability

All the data and code to reproduce this analysis can be found at https://osf.io/kzq69/.

## Graphical abstract or supplemental file

“If a COVID-19 vaccine is proven safe and effective and is available, **I will take it”**

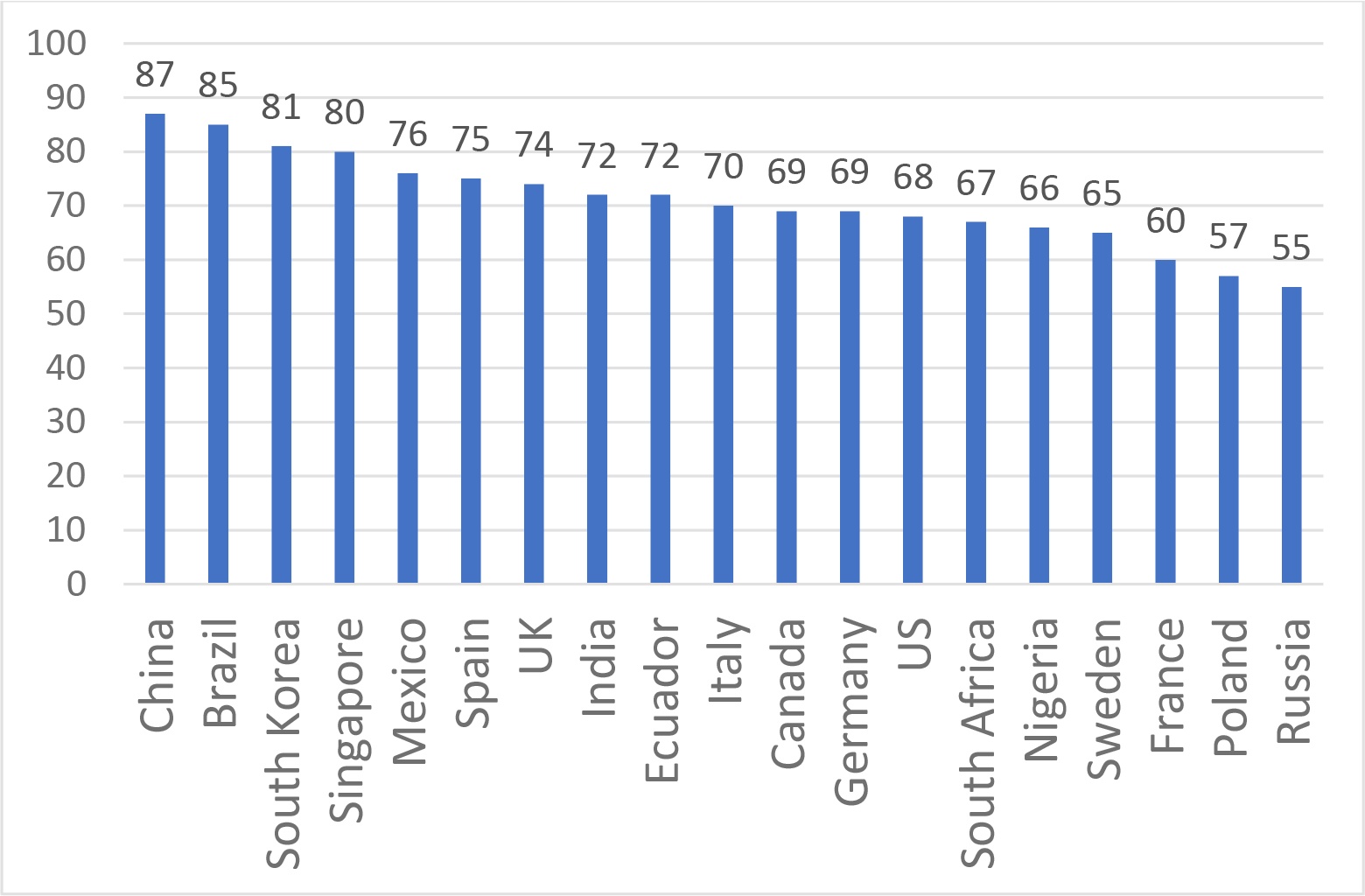

## Data sharing

All the data and code to reproduce this analysis can be found at https://osf.io/kzq69/.

## Acknowledgements

JVL acknowledges support to ISGlobal from the Spanish Ministry of Science, Innovation and Universities through the “Centro de Excelencia Severo Ochoa 2019–2023” Programme (CEX2018–000806-S) and from the Government of Catalonia through the CERCA Programme.

## Ethical approval

This study was approved by Emerson College, USA (IRB protocol number 20–023-F-E-6/12), with an expiration date of 11 June 2021. The online questionnaire was administered by Emerson College to gather information from respondents after obtaining their written, informed consent about the survey and this project. No personally identifiable information was collected or stored.

## Author contributions

SR, JVL and AEM conceived the study. SK collected the data. AP was responsible for the coding of the analyses. AP, AEM and JVL analysed the data. JVL, AP, KR and AEM wrote the first draft of the paper. JVL, AEM, AP, KR, SR, LOG, HL edited and approved the final manuscript.

## Declaration of interests

None declared for all authors. Dr Jeffrey V Lazarus affirms that the manuscript is an honest, accurate, and transparent account of the study being reported; that no important aspects of the study have been omitted. ICMJEs for all authors are available upon request.

**Supp Table 1:** Breakdown of demographic parameters and responses by country (n = 19)

|  | Brazil | Canada | China | Ecuador | France | Germany | India | Italy | Mexico | Nigeria | Poland | Russia | Singapore | South Africa | South Korea | Spain | Sweden | UK | US |
| --- | --- | --- | --- | --- | --- | --- | --- | --- | --- | --- | --- | --- | --- | --- | --- | --- | --- | --- | --- |
| N | 717 | 707 | 712 | 741 | 669 | 722 | 742 | 736 | 699 | 670 | 666 | 680 | 655 | 619 | 752 | 748 | 650 | 768 | 773 |
| Gender |  |  |  |  |  |  |  |  |  |  |  |  |  |  |  |  |  |  |  |
| Female | 436<br>(60.9) | 392<br>(55.6) | 351<br>(49.4) | 407<br>(55.0) | 333<br>(49.8) | 417<br>(58.2) | 485<br>(66.1) | 412<br>(56.0) | 364<br>(52.1) | 373<br>(55.7) | 302<br>(45.5) | 346<br>(50.9) | 310<br>(47.3) | 294<br>(47.6) | 392<br>(52.3) | 401<br>(53.6) | 326<br>(50.2) | 408<br>(53.3) | 423<br>(55.0) |
| Male | 276<br>(38.5) | 307<br>(43.5) | 360<br>(50.6) | 323<br>(43.6) | 334<br>(49.9) | 298<br>(41.6) | 243<br>(33.1) | 323<br>(43.9) | 332<br>(47.6) | 275<br>(41.0) | 362<br>(54.5) | 328<br>(48.2) | 342<br>(52.2) | 321<br>(51.9) | 357<br>(47.7) | 345<br>(46.1) | 322<br>(49.5) | 344<br>(44.9) | 337<br>(43.8) |
| Other | 4 (<br>0.6) | 6 (<br>0.9) | 0 (<br>0.0) | 10 (<br>1.4) | 2 (<br>0.3) | 2 ( 0.3) | 6 (<br>0.8) | 1 (<br>0.1) | 2 (<br>0.3) | 22 (<br>3.3) | 0 (<br>0.0) | 6 (<br>0.9) | 3 ( 0.5) | 3 (<br>0.5) | 0 (0.0) | 2 (<br>0.3) | 2 ( 0.3) | 14 (<br>1.8) | 9 (<br>1.2) |
| Income |  |  |  |  |  |  |  |  |  |  |  |  |  |  |  |  |  |  |  |
| <\$2 per day | 20 (<br>2.8) | 22 (<br>3.1) | 2 (<br>0.3) | 30 (<br>4.0) | 8 (<br>1.2) | 2 ( 0.3) | 39 (<br>5.3) | 4 (<br>0.5) | 15 (<br>2.1) | 195<br>(29.1) | 13 (<br>2.0) | 24 (<br>3.5) | 22 ( 3.4) | 9 (<br>1.5) | 4<br>(0.5) | 5 (<br>0.7) | 18 (<br>2.8) | 8 (<br>1.0) | 7 (<br>0.9) |
| \$2–\$8 per day | 89<br>(12.4) | 9 (<br>1.3) | 3 (<br>0.4) | 92<br>(12.4) | 5 (<br>0.7) | 5 ( 0.7) | 172<br>(23.2) | 2 (<br>0.3) | 88<br>(12.6) | 225<br>(33.6) | 5 (<br>0.8) | 42 (<br>6.2) | 37 ( 5.6) | 47 (<br>7.6) | 4<br>(0.5) | 2 (<br>0.3) | 1 ( 0.2) | 5 (<br>0.7) | 7 (<br>0.9) |
| \$8–\$32 per day | 334<br>(46.6) | 56 (<br>7.9) | 69 (<br>9.7) | 344<br>(46.4) | 48 (<br>7.2) | 51 ( 7.1) | 360<br>(48.5) | 44 (<br>6.0) | 306<br>(43.8) | 175<br>(26.1) | 316<br>(47.4) | 340<br>(50.0) | 236<br>(36.0) | 184<br>(29.7) | 42<br>(5.6) | 29 (<br>3.9) | 2 ( 0.3) | 55 (<br>7.2) | 20 (<br>2.6) |
| \$32+ | 238<br>(33.2) | 567<br>(80.2) | 633<br>(88.9) | 225<br>(30.4) | 561<br>(83.9) | 634<br>(87.8) | 163<br>(22.0) | 651<br>(88.5) | 263<br>(37.6) | 30 (<br>4.5) | 286<br>(42.9) | 256<br>(37.6) | 339<br>(51.8) | 329<br>(53.2) | 678<br>(90.2) | 692<br>(92.5) | 593<br>(91.2) | 644<br>(83.9) | 716<br>(92.6) |
| Refused | 36 (<br>5.0) | 53 (<br>7.5) | 5 (<br>0.7) | 50 (<br>6.7) | 47 (<br>7.0) | 30 ( 4.2) | 8 (<br>1.1) | 35 (<br>4.8) | 27 (<br>3.9) | 45 (<br>6.7) | 46 (<br>6.9) | 18 (<br>2.6) | 21 ( 3.2) | 50 (<br>8.1) | 24<br>(3.2) | 20 (<br>2.7) | 36 (<br>5.5) | 56 (<br>7.3) | 23 (<br>3.0) |
| Education |  |  |  |  |  |  |  |  |  |  |  |  |  |  |  |  |  |  |  |
| Less than high school | 176<br>(24.6) | 204<br>(28.9) | 236<br>(33.2) | 371<br>(50.1) | 407<br>(60.8) | 338<br>(47.1) | 126<br>(17.2) | 115<br>(15.6) | 207<br>(29.7) | 249<br>(37.2) | 61 (<br>9.2) | 67 (<br>9.9) | 205<br>(31.3) | 136<br>(22.0) | 183<br>(24.4) | 199<br>(26.6) | 325<br>(50.0) | 167<br>(21.8) | 58 (<br>7.5) |
| High school some college | 272<br>(38.0) | 380<br>(53.9) | 436<br>(61.3) | 276<br>(37.3) | 134<br>(20.0) | 133<br>(18.5) | 429<br>(58.4) | 378<br>(51.4) | 413<br>(59.2) | 325<br>(48.5) | 295<br>(44.4) | 157<br>(23.1) | 219<br>(33.4) | 85<br>(13.8) | 145<br>(19.4) | 129<br>(17.2) | 146<br>(22.5) | 197<br>(25.7) | 143<br>(18.6) |
| Bachelor | 232<br>(32.4) | 97<br>(13.8) | 33 (<br>4.6) | 77<br>(10.4) | 105<br>(15.7) | 118<br>(16.5) | 163<br>(22.2) | 107<br>(14.5) | 66 (<br>9.5) | 82<br>(12.2) | 308<br>(46.4) | 405<br>(59.6) | 178<br>(27.2) | 318<br>(51.5) | 340<br>(45.4) | 309<br>(41.3) | 114<br>(17.5) | 254<br>(33.2) | 388<br>(50.5) |
| Postgraduate | 36 (<br>5.0) | 24 (<br>3.4) | 6 (<br>0.8) | 16 (<br>2.2) | 23 (<br>3.4) | 128<br>(17.9) | 16 (<br>2.2) | 136<br>(18.5) | 12 (<br>1.7) | 14 (<br>2.1) | 0 (<br>0.0) | 51 (<br>7.5) | 53 ( 8.1) | 79<br>(12.8) | 81<br>(10.8) | 111<br>(14.8) | 65<br>(10.0) | 148<br>(19.3) | 180<br>(23.4) |
| Age |  |  |  |  |  |  |  |  |  |  |  |  |  |  |  |  |  |  |  |
| 18–24 | 171<br>(23.9) | 102<br>(14.5) | 85<br>(12.0) | 224<br>(30.3) | 105<br>(15.7) | 84<br>(11.7) | 58 (<br>7.9) | 111<br>(15.1) | 134<br>(19.2) | 203<br>(30.3) | 84<br>(12.7) | 76<br>(11.2) | 118<br>(18.0) | 77<br>(12.5) | 83<br>(11.1) | 115<br>(15.4) | 63 (<br>9.7) | 121<br>(15.8) | 43 (<br>5.6) |
| 25–54 | 412<br>(57.5) | 428<br>(60.7) | 450<br>(63.3) | 425<br>(57.4) | 381<br>(57.0) | 403<br>(56.2) | 599<br>(81.6) | 457<br>(62.1) | 416<br>(59.6) | 439<br>(65.5) | 382<br>(57.5) | 441<br>(64.9) | 399<br>(60.9) | 400<br>(64.7) | 499<br>(66.6) | 486<br>(65.0) | 297<br>(45.7) | 479<br>(62.5) | 567<br>(73.7) |
| 55–64 | 57 (8.0) | 86<br>(12.2) | 62 (8.7) | 58 (7.8) | 77<br>(11.5) | 117<br>(16.3) | 48 (6.5) | 94<br>(12.8) | 69 (9.9) | 28 (4.2) | 95<br>(14.3) | 86<br>(12.6) | 84 (12.8) | 70<br>(11.3) | 97<br>(13.0) | 70 (9.4) | 128<br>(19.7) | 79<br>(10.3) | 88<br>(11.4) |
| 65+ | 76<br>(10.6) | 89<br>(12.6) | 114<br>(16.0) | 33 (4.5) | 106<br>(15.8) | 113<br>(15.8) | 29 (4.0) | 74<br>(10.1) | 79<br>(11.3) | 0 (0.0) | 103<br>(15.5) | 77<br>(11.3) | 54 (8.2) | 71<br>(11.5) | 70<br>(9.3) | 77<br>(10.3) | 162<br>(24.9) | 87<br>(11.4) | 71 (9.2) |
| Vaccine question general |  |  |  |  |  |  |  |  |  |  |  |  |  |  |  |  |  |  |  |
| Completely agree | 481<br>(67.1) | 341<br>(48.2) | 413<br>(58.0) | 383<br>(51.7) | 202<br>(30.2) | 327<br>(45.3) | 329<br>(44.3) | 323<br>(43.9) | 395<br>(56.5) | 310<br>(46.3) | 241<br>(36.2) | 181<br>(26.6) | 319<br>(48.7) | 316<br>(51.1) | 327<br>(43.5) | 400<br>(53.5) | 296<br>(45.5) | 361<br>(47.0) | 341<br>(44.1) |
| Somewhat agree | 131<br>(18.3) | 145<br>(20.5) | 218<br>(30.6) | 150<br>(20.2) | 192<br>(28.7) | 167<br>(23.1) | 224<br>(30.2) | 198<br>(26.9) | 138<br>(19.7) | 127<br>(19.0) | 134<br>(20.1) | 192<br>(28.2) | 126<br>(19.2) | 189<br>(30.5) | 273<br>(36.3) | 156<br>(20.9) | 128<br>(19.7) | 188<br>(24.5) | 242<br>(31.3) |
| Neutral/no opinion | 57 (7.9) | 106<br>(15.0) | 76<br>(10.7) | 94<br>(12.7) | 152<br>(22.7) | 111<br>(15.4) | 105<br>(14.2) | 109<br>(14.8) | 83<br>(11.9) | 94<br>(14.0) | 109<br>(16.4) | 128<br>(18.8) | 73 (11.1) | 84<br>(13.6) | 114<br>(15.2) | 92<br>(12.3) | 117<br>(18.0) | 105<br>(13.7) | 103<br>(13.3) |
| Somewhat disagree | 29 (4.0) | 55 (7.8) | 5 (0.7) | 40 (5.4) | 49 (7.3) | 43 (6.0) | 42 (5.7) | 51 (6.9) | 29 (4.1) | 53 (7.9) | 64 (9.6) | 75<br>(11.0) | 40 (6.1) | 21<br>(3.4) | 27<br>(3.6) | 53 (7.1) | 56 (8.6) | 54 (7.0) | 33 (4.3) |
| Completely disagree | 19 (2.6) | 60 (8.5) | 0 (0.0) | 74<br>(10.0) | 74<br>(11.1) | 74<br>(10.2) | 42 (5.7) | 55 (7.5) | 54 (7.7) | 86<br>(12.8) | 118<br>(17.7) | 104<br>(15.3) | 97 (14.8) | 9 (1.5) | 11<br>(1.5) | 47 (6.3) | 53 (8.2) | 60 (7.8) | 54 (7.0) |
| Vaccine question emplyer |  |  |  |  |  |  |  |  |  |  |  |  |  |  |  |  |  |  |  |
| Completely agree | 68 (9.5) | 78<br>(11.0) | 306<br>(43.0) | 48 (6.5) | 49 (7.3) | 118<br>(16.3) | 184<br>(24.8) | 55 (7.5) | 78<br>(11.2) | 96<br>(14.3) | 78<br>(11.7) | 37 (5.4) | 97 (14.8) | 74<br>(12.0) | 174<br>(23.1) | 75<br>(10.0) | 65<br>(10.0) | 92<br>(12.0) | 109<br>(14.1) |
| Somewhat agree | 190<br>(26.5) | 280<br>(39.6) | 290<br>(40.7) | 153<br>(20.6) | 239<br>(35.7) | 271<br>(37.5) | 290<br>(39.1) | 300<br>(40.8) | 233<br>(33.3) | 200<br>(29.9) | 215<br>(32.3) | 147<br>(21.6) | 245<br>(37.4) | 208<br>(33.6) | 362<br>(48.1) | 234<br>(31.3) | 162<br>(24.9) | 271<br>(35.3) | 289<br>(37.4) |
| Neutral/no opinion | 158<br>(22.0) | 227<br>(32.1) | 90<br>(12.6) | 192<br>(25.9) | 197<br>(29.4) | 209<br>(28.9) | 125<br>(16.8) | 220<br>(29.9) | 157<br>(22.5) | 158<br>(23.6) | 161<br>(24.2) | 218<br>(32.1) | 153<br>(23.4) | 211<br>(34.1) | 161<br>(21.4) | 178<br>(23.8) | 257<br>(39.5) | 234<br>(30.5) | 182<br>(23.5) |
| Somewhat disagree | 206<br>(28.7) | 89<br>(12.6) | 23 (3.2) | 193<br>(26.0) | 135<br>(20.2) | 92<br>(12.7) | 84<br>(11.3) | 115<br>(15.6) | 146<br>(20.9) | 130<br>(19.4) | 134<br>(20.1) | 184<br>(27.1) | 120<br>(18.3) | 90<br>(14.5) | 35<br>(4.7) | 173<br>(23.1) | 121<br>(18.6) | 104<br>(13.5) | 125<br>(16.2) |
| Completely disagree | 95<br>(13.2) | 33 (4.7) | 3 (0.4) | 155<br>(20.9) | 49 (7.3) | 32 (4.4) | 59 (8.0) | 46 (6.2) | 85<br>(12.2) | 86<br>(12.8) | 78<br>(11.7) | 94<br>(13.8) | 40 (6.1) | 36 (5.8) | 20<br>(2.7) | 88<br>(11.8) | 45 (6.9) | 67 (8.7) | 68 (8.8) |

